# Neurodevelopmental outcomes of infants secondary to in utero exposure to maternal SARS-CoV-2 infection: A national prospective study in Kuwait

**DOI:** 10.1101/2021.11.12.21266291

**Authors:** Mariam Ayed, Alia Embaireeg, Mais Kartam, Kiran More, Mafaza Alqallaf, Abdullah AlNafisi, Zainab Alsaffar, Zainab Bahzad, Yasmeen Buhamad, Haneen Alsayegh, Wadha Al-Fouzan, Hessa Alkandari

**Author notes:** **CORRESPONDING AUTHOR:** Mariam Ayed, Neonatal Department, Farwaniya Hospital, Subah An Nasser, Kuwait City, Postal code-81400, Kuwait.

## Abstract

**Background:** An increasing proportion of women are being infected with severe acute respiratory syndrome coronavirus 2 (SARS-CoV-2) during pregnancy. Intrauterine viral infections induce an increase in the levels of proinflammatory cytokines, which inhibit the proliferation of neuronal precursor cells and stimulate oligodendrocyte cell death, leading to abnormal neurodevelopment. Whether a maternal cytokine storm can affect neonatal brain development is unclear. The objective of the present study is to assess neurodevelopmental outcomes in neonates born to mothers with SARS-CoV-2 infections during pregnancy.

**Methods:** In this prospective cohort study, the neurodevelopment status of infants (N=298) born to women with SARS-CoV-2 infections during pregnancy was assessed at 10-12 months post discharge using the Ages and Stages Questionnaire, 3rd edition (ASQ-3). The ASQ-3 scores were classified into developmental delays (cutoff score: ≤2 standard deviations (SDs) below the population mean) and no delay (score >2 SDs above the population mean).

**Results:** Approximately 10% of infants born to mothers with SARS-CoV-2 infections during pregnancy showed developmental delays. Two of 298 infants tested positive for SARS-CoV-2, and both had normal ASQ-3 scores. The majority of the pregnant women had SARS-CoV-2 infection during their third trimester. The risk of developmental delays among infants was higher in those whose mothers had SARS-CoV-2 infections during the first (P=0.039) and second trimesters (P=0.001) than in those whose mothers had SARS-CoV-2 infections during the third trimester. Infants born at <31 weeks gestation were more prone to developmental delays than those born at >31 weeks gestation (10% versus 0.8%; P=0.002).

**Conclusion:** The findings of the study highlight the need for long term neurodevelopmental assessment of infants born to mothers with SARS-CoV-2 infection.

## Background

Severe acute respiratory syndrome coronavirus 2 (SARS-CoV-2), which causes coronavirus disease-19 (COVID-19), has been spreading rapidly worldwide, increasingly affecting even pregnant females. Pregnancy-associated physiological changes, as well as altered cell-mediated immunity, enhance the susceptibility of pregnant women to infections by intracellular organisms such as viruses. Based on a living systematic review and meta-analysis, overall, 10% (95% confidence interval (CI): 7-12%; 73 studies involving 67271 women) of pregnant and recently pregnant women attending or admitted to the hospital for any reason were diagnosed as having suspected or confirmed SARS-CoV-2 infection [**1**]. Hence, a growing proportion of pregnant women become infected with SARS-CoV-2 during various trimesters of pregnancy, with variable effects on the fetus [**1,2**].

Furthermore, the underdeveloped innate and adaptive immune systems of fetuses and neonates also make them more susceptible to infections **[3, 4]**. Maternal-fetal transmission of SARS-CoV-2 has not been widely established to date, and findings vary. A meta-analysis revealed that vertical transmission of SARS-CoV-2 is possible and showed that only 3.2% (27 of 936; 95% CI: 2.2-4.3) of neonates from mothers infected with SARS-CoV-2 had a positive result based on viral RNA testing of nasopharyngeal swabs **[5]**. SARS-CoV-2 viral RNA testing was positive in neonatal cord blood (2.9% samples), placental swabs (7.7%), and fecal or rectal swabs, whereas it was negative in urine and amniotic fluid samples. In contrast, a more recent systematic review article (2021), which covered the database up to September 2020, reported that newborn rates of the infection vary between 0% and 11.5% [**6**]. Nevertheless, due to the availability of only a few patients and studies, the exact rates of vertical transmission and fetal or neonatal morbidity and mortality cannot be ascertained.

Although SARS-CoV-2 primarily causes respiratory distress, it is also known to have extrapulmonary manifestations, including hematological, cardiovascular, endocrinological, and neurological complications, in the adult population **[7-9]**. Of these, neurological manifestations in pregnant women and their probable effect on fetuses and neonates are of primary concern. SARS-CoV-2 may gain entry into the central nervous system through the nasal mucosa, lamina cribrosa, and olfactory bulb or through retrograde axonal transport. SARS-CoV-2 also exhibits neurovirulence, triggering proinflammatory and prothrombotic cascades in the wake of cytokine storms, affecting brain vasculature, as well as the blood-brain barrier, mainly in the setting of the toxic-metabolic sequelae of multiorgan dysfunction frequently observed in SARS-CoV-2-positive patients **[10,11]**. Proinflammatory cytokines inhibit the proliferation of neuronal precursor cells, activate astrogliosis, and stimulate oligodendrocyte cell death, leading to abnormal neurodevelopment **[3, 4]**. Moreover, the elevated levels of maternal inflammatory cytokines (IL-1, IL-6, IL-8, and TNF-α) as a consequence of infection during pregnancy can disturb several characteristics of fetal brain development [**12**]. Notably, these pathological changes, which include deteriorated neuronal functions and atypical behavioral changes, can subsequently be seen in postnatal life and may increase the risk of schizophrenia, autism, and mental disorders **[12, 13]**.

Additionally, animal studies have shown that offspring from maternal immune activation animal models have schizophrenia and autism-related behaviors, including decreased sensorimotor gating, deficits in working memory and cognitive flexibility, increased anxiety, and enhanced sensitivity to amphetamines **[14-16]**.

Because of the plausible effects on neonates born to mothers with SARS-CoV-2 infections during pregnancy, this study assessed the developmental attainment at 10-12 months of neonates born to a cohort of mothers with SARS-CoV-2 infections during pregnancy in Kuwait using the Ages and Stages Questionnaire, 3rd edition (ASQ-3).

## 2 Methods

### 2.1 Participants

In this prospective cohort study, infants born between April 1 and August 30, 2020, to mothers with SARS-CoV-2 infection during various trimesters of pregnancy were evaluated. Pregnant women who tested positive by reverse transcription-polymerase chain reaction (RT-PCR) for SARS-CoV-2 (Cobas 6800 Systems, Roche, Switzerland)/(TaqPath, Thermo-Fisher Scientific, USA) were identified from the Kuwait National COVID-19 registry. Mothers with equivocal RT-PCR test results and those with missing data were not included in the study. Verbal informed consent was obtained from all the parents who participated in the study. Ethics approval was granted by the Ministry of Health, (2021-1638), Government of Kuwait.

Demographic details were collected for all infants enrolled in the follow-up study. Maternal clinical and neonatal data from acute hospital admissions were collected by retrospective chart review of the included participants and have been previously published **[17]**.

### 2.2 Instrument

The ASQ-3 (http://agesandstages.com/) is a widely used screening tool for assessing development in children aged 1–66 months at a low cost, with cutoff scores identifying developmental delays. The screening tool has strong validity, and its use has been widespread in assessing development in children **[18]**. The ASQ-3 has five domains: communication, gross motor skills, fine motor skills, problem-solving capacity, and personal-social development.

In the current study, we used the ASQ-3 to assess neurodevelopmental outcomes at 10-12 months postdischarge. Parents of infants completed the ASQ-3, which has a series of 21 developmental screening questions. Three study investigators (MAL, MK, and AE) scored the questionnaire results using the ASQ-3 age-specific scoring sheet. In each domain, six items were scored as present in the infant, sometimes present or not yet shown by the infant, with 10, 5, and 0 points, respectively. If an item was not filled in, a score of 0 was assigned based on the lowest possible result. Responses were summed to give a score of 0 to 60 for each domain and an overall maximum ASQ-3 score of 300 points. ASQ-3 results were categorized according to questionnaire-defined subscale cutoff scores into 1) developmental delays, with an ASQ-3 cutoff score of ≤2 standard deviations (SDs) below the population mean on 1 or more domains, and 2) no delay, with a cutoff score >2 SDs above the population mean **[18]**.

The questionnaire was sent in two languages: the Arabic version for Arabic-speaking parents and the English version for non-Arabic-speaking parents. The questionnaire was completed by parents and reviewed in a telephone interview with one investigator. Parents were encouraged to provide their responses with their interpretation without prompting.

### 2.3 Statistical analysis

All data were entered into a Microsoft Excel database. Categorical variables were summarized as counts (n) and percentages (%), and continuous variables were summarized as the median and interquartile range (IQR). Statistical comparisons between the groups were performed using the chi-squared test for categorical variables and the Wilcoxon rank-sum test for continuous variables.

We performed a multivariate logistic regression analysis for each potential predictor of developmental delays. Variables with a P value of <0.1 on the univariate analysis were included in the model (maternal age and trimester at SARS-CoV-2 infection) and adjusted for gestational age, birth weight, sex, parental education and type of feeding in the first 6 months of age. The results of the regression analysis are presented as an adjusted odds ratio (aOR) with a 95% CI. Statistical significance was taken at P<0.05. Statistical analysis was performed with **STATA 14** software (**Stata** Corporation, College Station, TX).

## 3 Results

Eight hundred forty pregnant women were identified from the national COVID-19 registry between April 1 and August 30, 2020. Four hundred forty-five women gave birth during the study period **(Figure 1)**. One infant died on the first day of life due to severe birth asphyxia; 298 (67%) had complete follow-up at 10-12 months corrected age, and 146 (32.8%) infants were lost to follow-up. The main reason for the loss to follow-up was the inability to contact the parents due to either invalid contact information or parents having left the country. To determine whether the infants we lost to follow-up were representative of the complete study and to avoid attrition bias, we compared the baseline features of those infants with follow-up data with the baseline features of infants who were lost to follow-up. We did not find any statistically significant differences between these two groups.

**Figure 1:**
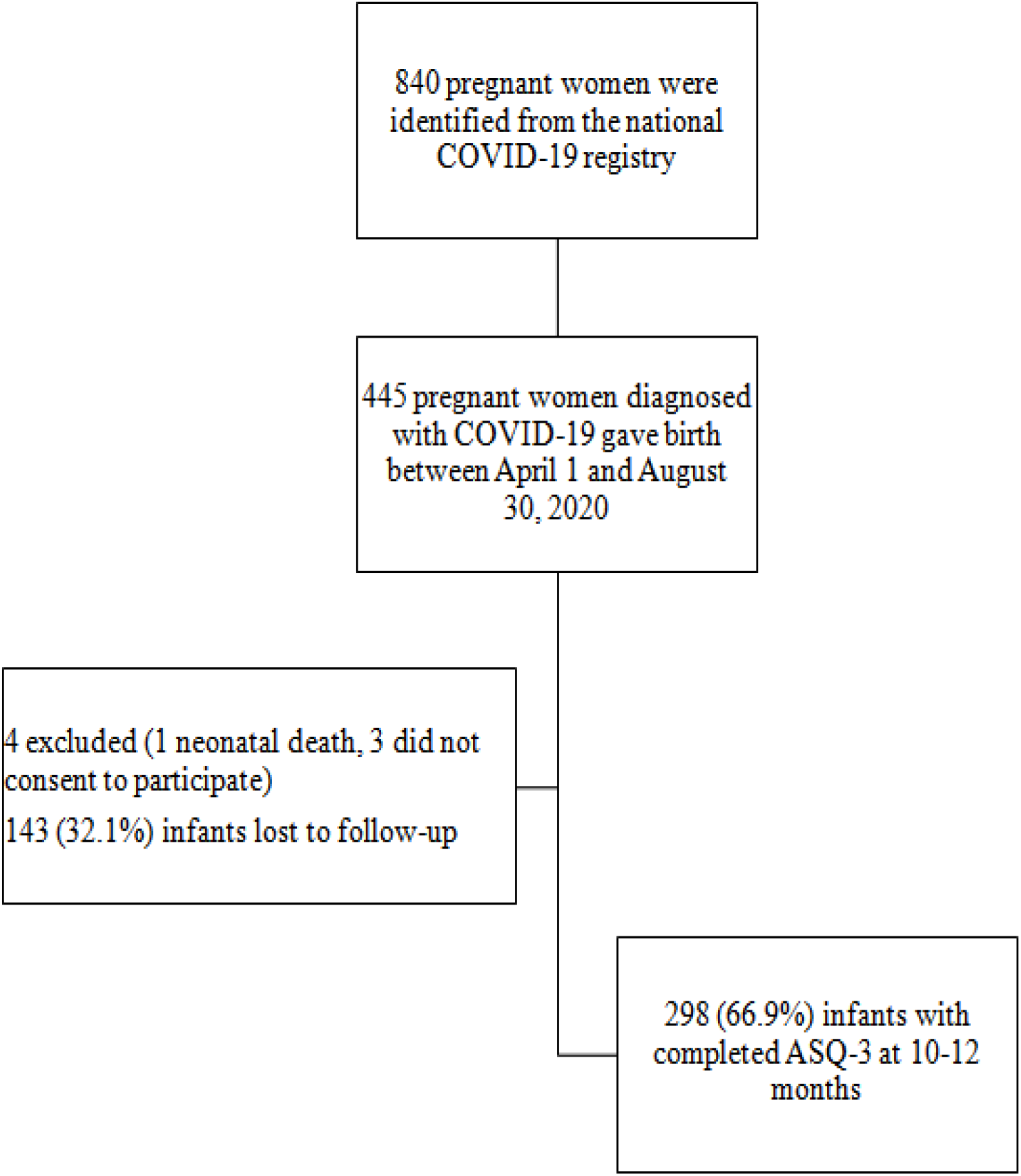
Flowchart of the study recruitment and follow-up. ASQ-3: Ages and Stages Questionnaire, 3rd edition.

The maternal and neonatal characteristics of the cohort are shown in **Table 1**.

**Table 1:**
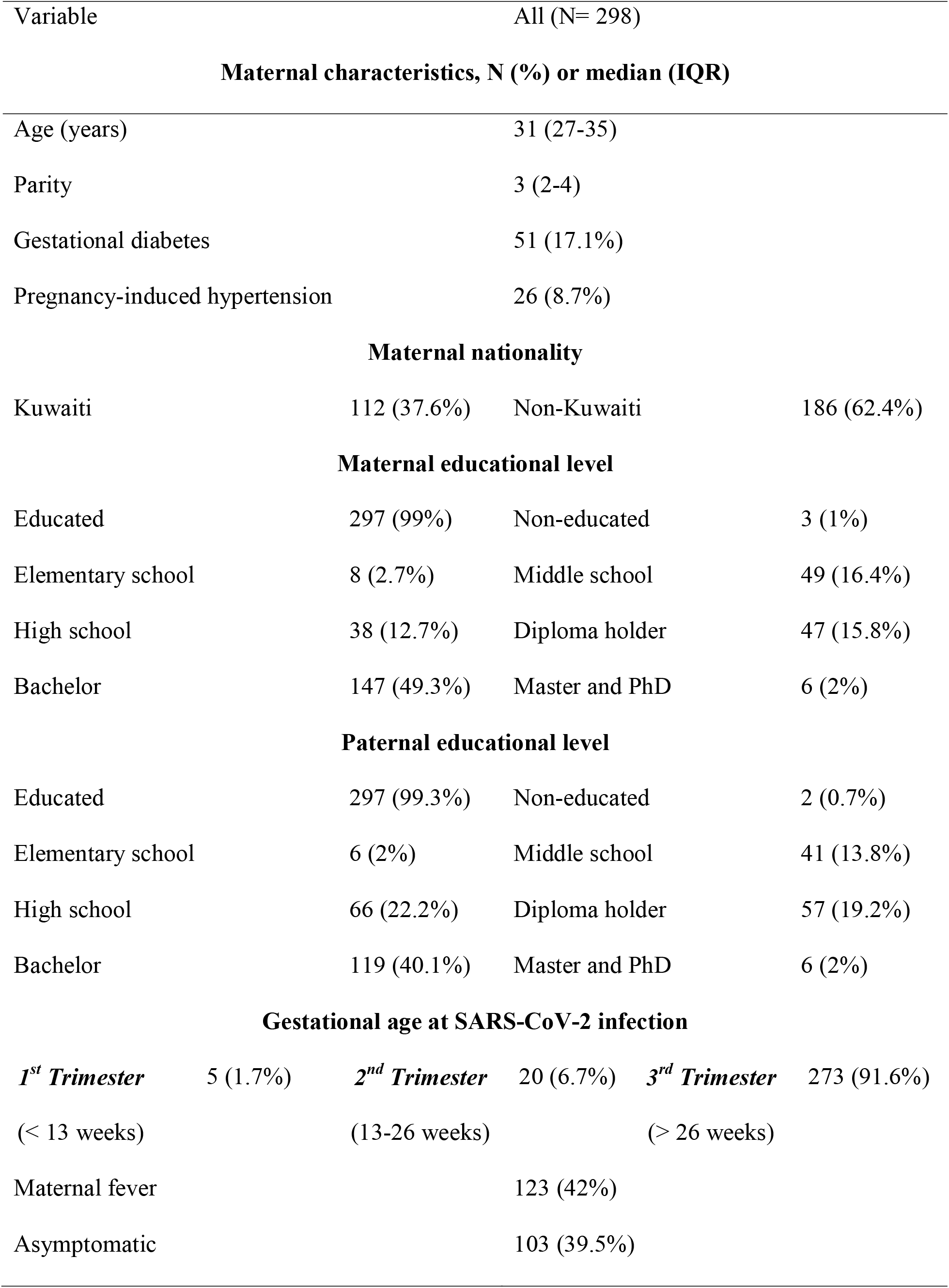
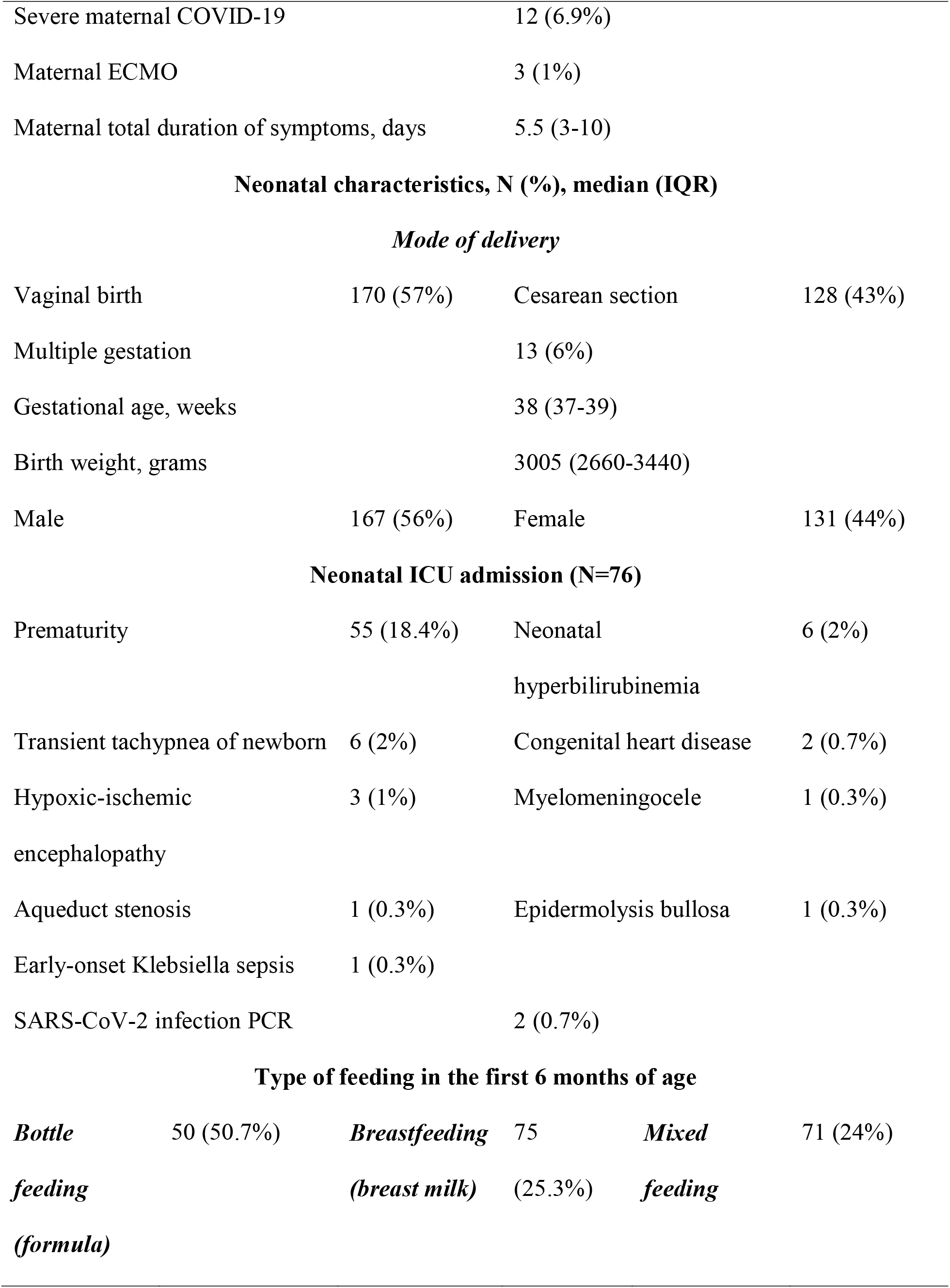
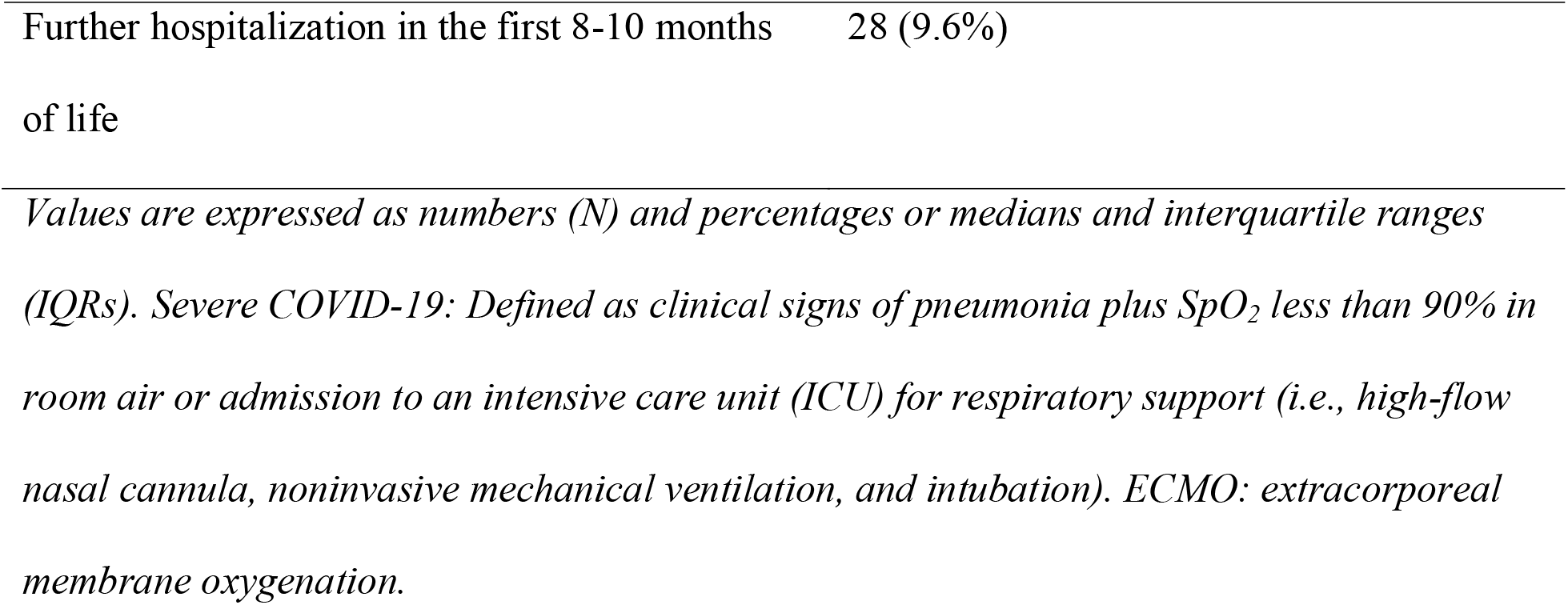
Maternal and neonatal demographic and clinical characteristics

Among the infants with completed ASQ-3, 5 (1.7%) were born to mothers diagnosed with SARS-CoV-2 during the first trimester, 20 (6.7%) during the second trimester, and 273 (91.6%) during the third trimester. Only 2 (0.7%) neonates tested positive for SARS-CoV-2 infection. Twenty-eight infants needed hospitalization beyond the first 8-10 months of age.

Parental questionnaires were collected at a median of 10.3 months corrected age (IQR 10 to 12.7). Developmental delays were identified in 30 (10.1%) of the infants. Of those with developmental delays, 3.3% (1/30) had delays in the communication domains, 27% (8/30) in the gross motor domain, 40% (12/30) in the fine motor domain, 10% (3/30) in the problem-solving domain, and 30% (9/30) in the personal-social domain **(Figure 2)**.

**Figure 2:**
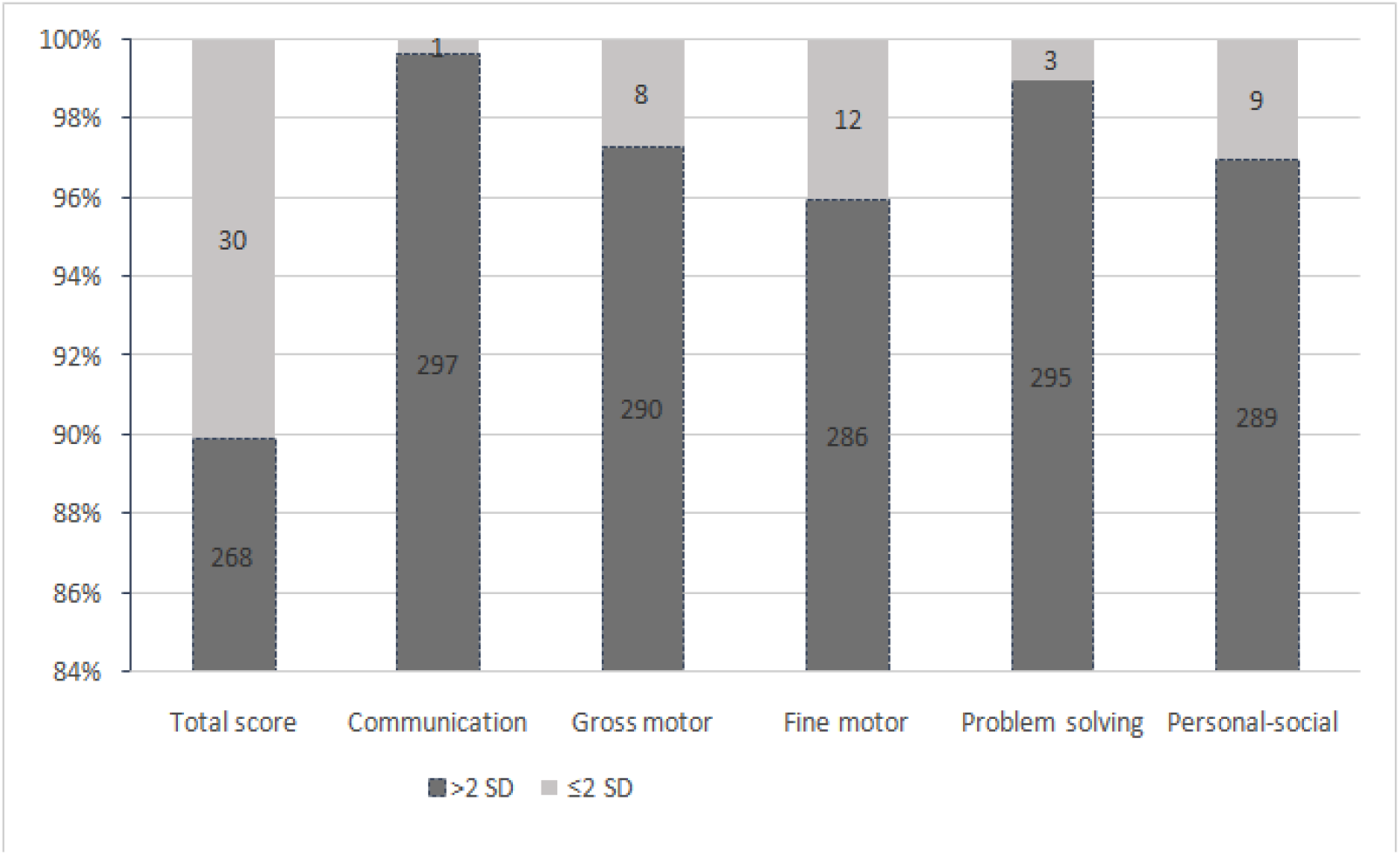
Developmental outcome using the Ages and Stages Questionnaire, 3rd edition, at 10-12 months of age. A bar graph presenting the total score and subscale scores of ASQ-3 domains. A score with a standard deviation (SD) ≤ 2 below the population mean implies developmental delays, and a score with an SD >2 above the population mean is considered no delay.

In the infants with developmental delays, 4 had concerns in two or more domains, and none had concerns in all domains.

Infants with developmental delays did not differ in terms of maternal age, maternal comorbidities, parental education level, severe maternal COVID-19, or the mode of delivery compared to those with no delay (**Table 2**). However, the occurrence of developmental delays differed significantly on the basis of the trimester at SARS-CoV-2 infection. Developmental delays were more common in infants born to mothers with COVID-19 during the first and second trimesters than in infants born to mothers with COVID-19 during the third trimester (P<0.001). Among infants with developmental delays, 4 (13.3%) were born to mothers with SARS-CoV-2 infection during the first trimester, 6 (20%) were born to mothers with SARS-CoV-2 infection during the second trimester, and 20 (66.7%) were born to mothers with SARS-CoV-2 infection in the third trimester. One (0.4%) of the infants without delay was born to a mother with SARS-CoV-2 infection in the first trimester, 14 (5.2%) were born to mothers with SARS-CoV-2 infections in the second trimester, and 253 (94.4%) were born to mothers with SARS-CoV-2 infections in the third trimester. Moreover, infants born at less than 31 weeks gestation were more likely to have developmental delays than infants born at more than 31 weeks gestation (10% versus 0.8%; P=0.002). Neonatal characteristics were similar between the groups.

**Table 2:**
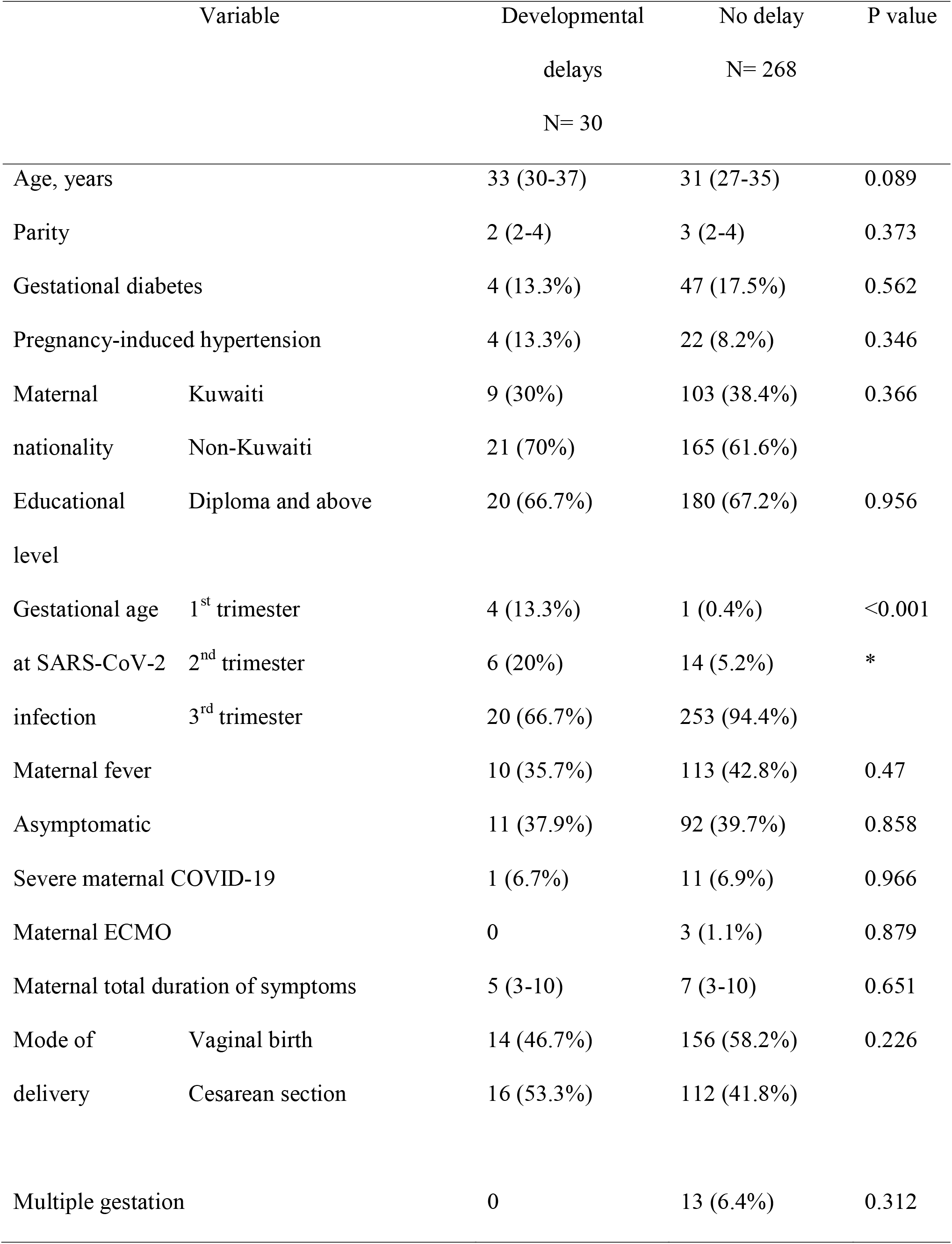
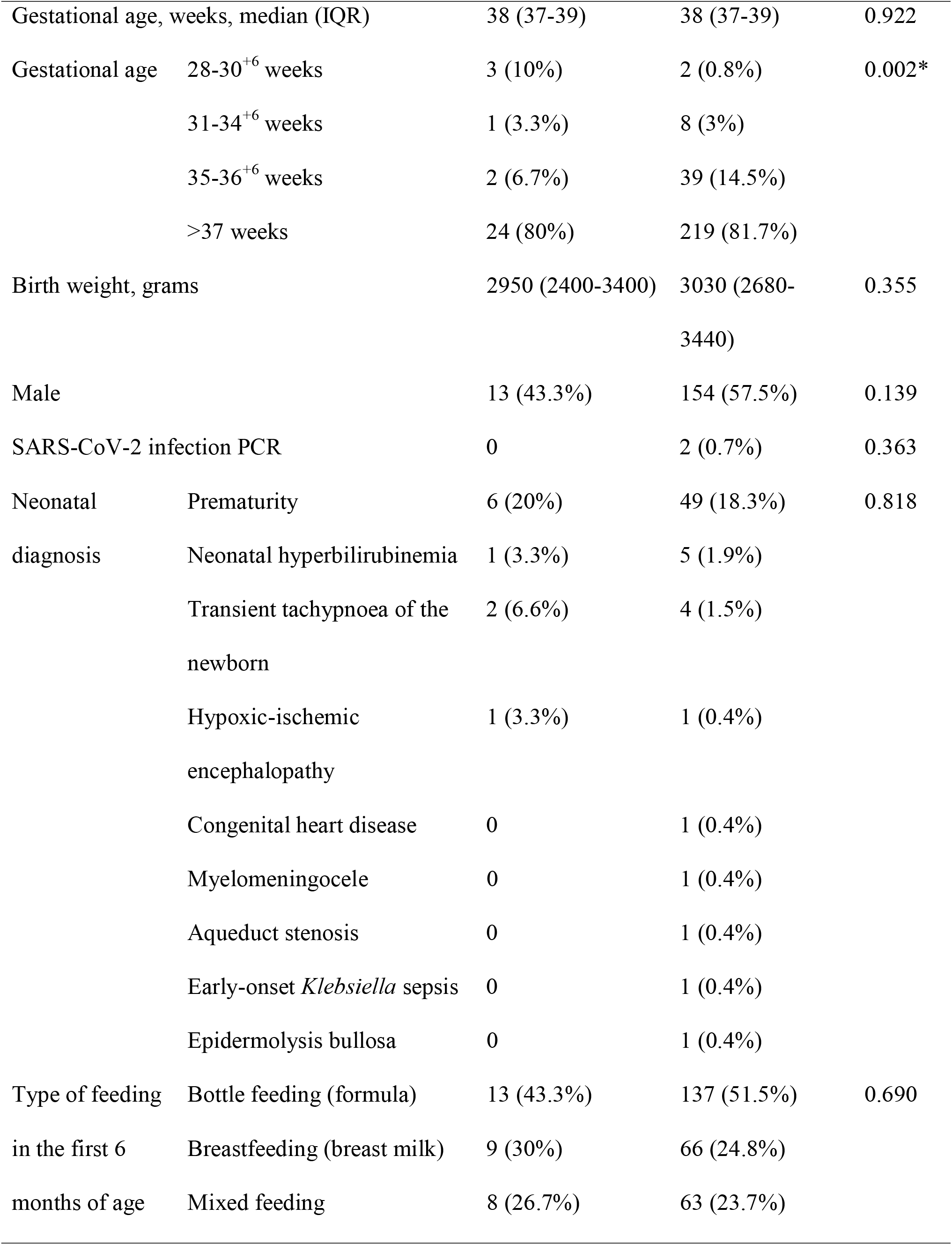
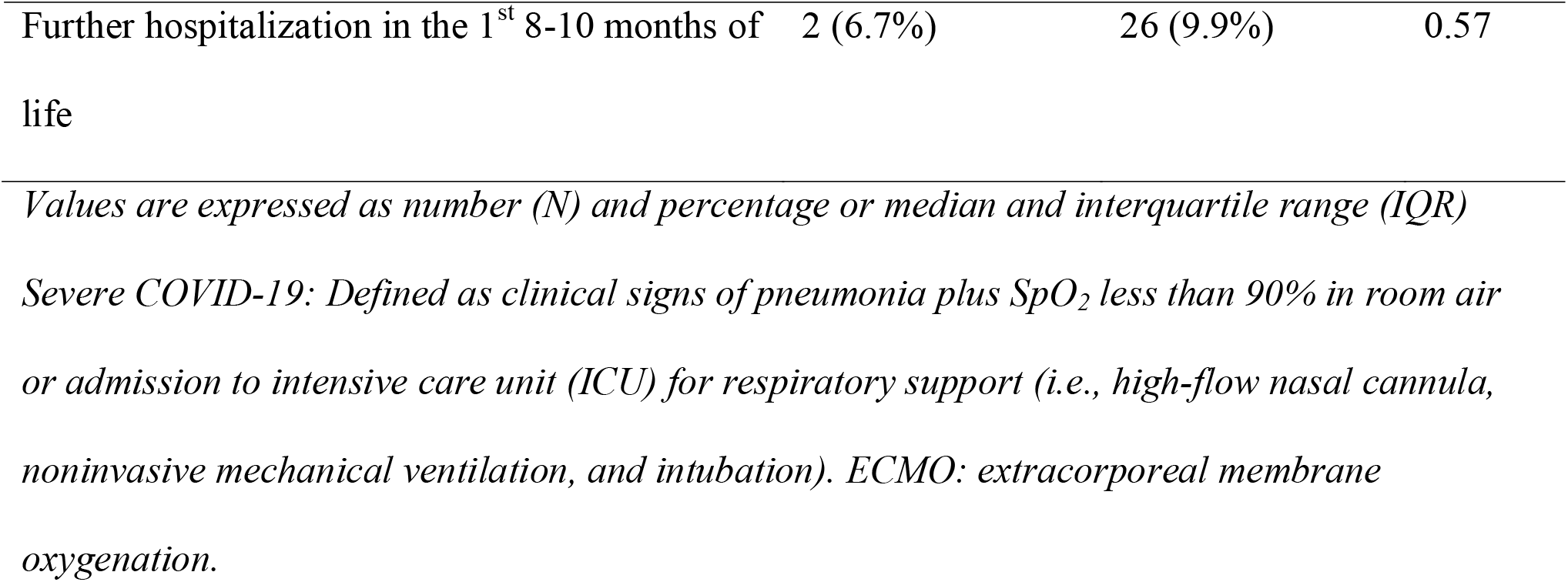
Maternal and neonatal demographic and clinical characteristics by the ASQ-3 results

| Variable |  | Developmental<br>delays<br>N= 30 | No delay<br>N= 268 | P value |
| --- | --- | --- | --- | --- |
| Age, years |  | 33 (30-37) | 31 (27-35) | 0.089 |
| Parity |  | 2 (2-4) | 3 (2-4) | 0.373 |
| Gestational diabetes |  | 4 (13.3%) | 47 (17.5%) | 0.562 |
| Pregnancy-induced hypertension |  | 4 (13.3%) | 22 (8.2%) | 0.346 |
| Maternal | Kuwaiti | 9 (30%) | 103 (38.4%) | 0.366 |
| nationality | Non-Kuwaiti | 21 (70%) | 165 (61.6%) |  |
| Educational level | Diploma and above | 20 (66.7%) | 180 (67.2%) | 0.956 |
| Gestational age at SARS-CoV-2 infection | 1<sup>st</sup> trimester | 4 (13.3%) | 1 (0.4%) | <0.001 |
|  | 2<sup>nd</sup> trimester | 6 (20%) | 14 (5.2%) | * |
|  | 3<sup>rd</sup> trimester | 20 (66.7%) | 253 (94.4%) |  |
| Maternal fever |  | 10 (35.7%) | 113 (42.8%) | 0.47 |
| Asymptomatic |  | 11 (37.9%) | 92 (39.7%) | 0.858 |
| Severe maternal COVID-19 |  | 1 (6.7%) | 11 (6.9%) | 0.966 |
| Maternal ECMO |  | 0 | 3 (1.1%) | 0.879 |
| Maternal total duration of symptoms |  | 5 (3-10) | 7 (3-10) | 0.651 |
| Mode of delivery | Vaginal birth | 14 (46.7%) | 156 (58.2%) | 0.226 |
|  | Cesarean section | 16 (53.3%) | 112 (41.8%) |  |
| Multiple gestation |  | 0 | 13 (6.4%) | 0.312 |
| Gestational age, weeks, median (IQR) |  | 38 (37-39) | 38 (37-39) | 0.922 |
| Gestational age | 28-30<sup>+6</sup> weeks | 3 (10%) | 2 (0.8%) | 0.002* |
|  | 31-34<sup>+6</sup> weeks | 1 (3.3%) | 8 (3%) |  |
|  | 35-36<sup>+6</sup> weeks | 2 (6.7%) | 39 (14.5%) |  |
|  | >37 weeks | 24 (80%) | 219 (81.7%) |  |
| Birth weight, grams |  | 2950 (2400-3400) | 3030 (2680-3440) | 0.355 |
| Male |  | 13 (43.3%) | 154 (57.5%) | 0.139 |
| SARS-CoV-2 infection PCR |  | 0 | 2 (0.7%) | 0.363 |
| Neonatal diagnosis | Prematurity | 6 (20%) | 49 (18.3%) | 0.818 |
|  | Neonatal hyperbilirubinemia | 1 (3.3%) | 5 (1.9%) |  |
|  | Transient tachypnoea of the newborn | 2 (6.6%) | 4 (1.5%) |  |
|  | Hypoxic-ischemic encephalopathy | 1 (3.3%) | 1 (0.4%) |  |
|  | Congenital heart disease | 0 | 1 (0.4%) |  |
|  | Myelomeningocele | 0 | 1 (0.4%) |  |
|  | Aqueduct stenosis | 0 | 1 (0.4%) |  |
|  | Early-onset *Klebsiella* sepsis | 0 | 1 (0.4%) |  |
|  | Epidermolysis bullosa | 0 | 1 (0.4%) |  |
| Type of feeding in the first 6 months of age | Bottle feeding (formula) | 13 (43.3%) | 137 (51.5%) | 0.690 |
|  | Breastfeeding (breast milk) | 9 (30%) | 66 (24.8%) |  |
|  | Mixed feeding | 8 (26.7%) | 63 (23.7%) |  |

|  |  |  |  |
| --- | --- | --- | --- |
| Further hospitalization in the 1<sup>st</sup> 8-10 months of life | 2 (6.7%) | 26 (9.9%) | 0.57 |

*Values are expressed as number (N) and percentage or median and interquartile range (IQR)*
*Severe COVID-19: Defined as clinical signs of pneumonia plus SpO<sub>2</sub> less than 90% in room air or admission to intensive care unit (ICU) for respiratory support (i.e., high-flow nasal cannula, noninvasive mechanical ventilation, and intubation). ECMO: extracorporeal membrane oxygenation.*

In multivariate logistic regression analysis, adjusting for maternal age, parental educational level, gestational age, birth weight, and the type of feeding in the first six months of age, the risk of developmental delays was higher with first-trimester maternal SARS-CoV-2 infection (aOR: 8.2, 95% CI: 1.1-55.9; P=0.039) and second-trimester maternal SARS-CoV-2 infection (aOR: 8.1, 95% CI: 2.4-27.7; P=0.001) than with third-trimester maternal SARS-CoV-2 infection **(Table 3)**.

**Table 3:** Multiple regression analysis of the association of demographic and clinical characteristics with developmental delays (ASQ-3 score less than 2 standard deviations below the population mean)

| Variable |  | Adjusted odds ratio | 95% CI | P value |
| --- | --- | --- | --- | --- |
| Trimester at | 3<sup>rd</sup> | Reference |  |  |
| SARS-CoV-2 | 2<sup>nd</sup> | 8.1 | 2.4-27.7 | 0.001* |
| infection | 1<sup>st</sup> | 8.2 | 1.1-55.9 | 0.039* |
| Maternal age |  | 1.04 | 0.96-1.1 | 0.305 |
| Gestational age at | 28-30<sup>+6</sup> weeks | 7.7 | 0.69-85.8 | 0.090 |
| birth | 31-34<sup>+6</sup> | 1.2 | 0.13-10.7 | 0.870 |
|  | 35-36<sup>+6</sup> | 0.5 | 0.10-2.3 | 0.371 |
|  | >37 | Reference |  |  |
| Maternal education (diploma and above) |  | 1.8 | 0.59-5.4 | 0.300 |
| Father education (diploma and above) |  | 0.5 | 0.2-1.3 | 0.172 |
| Male |  | 1.6 | 0.68-3.5 | 0.297 |
| Type of feeding | Mixed feeding | Reference |  |  |
| in the first 6 | Bottle feeding | 1.3 | 0.46-3.4 | 0.652 |
| months | (formula) |  |  |  |
|  | Breastfeeding | 0.5 | 0.14-1.6 | 0.239 |
|  | (breast milk) |  |  |  |

*Values are expressed as adjusted odds ratios and 95% confidence intervals (95% CIs).*

## 4 Discussions

The fetal inflammatory response (FIRS) due to maternal SARS-CoV-2 infection can contribute to severe neonatal morbidity, which includes stillbirth, neonatal death, preterm birth, low birth weight, fetal distress, and neonatal asphyxia [**19-21]**. This is perhaps the first study reporting neurological assessments of neonates born to mothers from West Asia with SARS-CoV-2-positive RT-PCR test results during pregnancy using the ASQ-3. The findings of this study indicate that infants born to mothers who test positive for SARS-CoV-2 infection during pregnancy need to be assessed for neurodevelopmental delays.

To assess the developmental attainment of infants born to mothers with SARS-CoV-2-positive test results during pregnancy, this study employed the ASQ-3, a screening tool used for assessing the developmental attainment of infants and young children **[18]**. This screening tool is used widely in assessing development in children aged 1–66 months at a low cost, with cutoff scores identifying developmental delays. In the current study, an Arabic version of the ASQ-3 was used for mothers who were well versed in Arabic. Since the study cohort included diverse as well as immigrant mothers, questionnaires were sent to mothers, and fortunately, the majority of them consented to respond.

The study data revealed that the majority (91.6%) of the pregnant women had SARS-CoV-2 infection during their third trimester, whereas 6.7% had SARS-CoV-2 infection in their second trimester. This finding is in agreement with those of other studies that have also reported that most SARS-CoV-2 infections in pregnant women occur in their third trimester **[22, 23]**. In the current study, only a relatively small number of women were infected during the first trimester. However, first-trimester infections are important as the principal stages of brain development, such as primary neurulation (weeks 3–4), prosencephalic development (months 2–3), and neuronal proliferation (months 3–4), occur during the early stages of pregnancy [**24**]. In fact, infections with some common pathogens, such as cytomegalovirus (CMV), Zika virus, Rubella virus, *Mycobacterium tuberculosis* (TB), and *Toxoplasma gondii*, during the first and early second trimesters increase the risk of symptomatic infants, with up to 32% being associated with neurological manifestations [**25]**. Owing to the relatively recent emergence of COVID-19, information related to pregnancy outcomes among women with SARS-CoV-2 infection and the consequences of infant exposure to the virus is very scarce. As more women become infected during their first and second trimesters progress in their pregnancy and as newborns with SARS-CoV-2 infection develop, a better understanding of the neurological effects of this novel virus will emerge. In brief, various types of evidence support the theory that maternal infection and/or inflammation occurring during critical periods of fetal development could alter brain structure and function in a time-sensitive manner.

Nevertheless, SARS-CoV-2 infection increases the chances of fetal distress, leading to high incidences of admission to the neonatal intensive care unit (NICU) [**23**]. Similar to other reported studies, in the current study, 76 out of 298 (25.5%) neonates born to mothers with SARS-CoV-2-infection during pregnancy required NICU admission [**26-29**]. On the other hand, a systematic review reported that approximately 95% of neonates born to mothers with SARS-CoV-2 infection during pregnancy were born in good condition [**1**].

Recent evidence suggests that vertical transmission of SARS-CoV-2 either antenatally or intrapartum can occur, but it is uncommon. In this study, only 2 (0.7%) neonates tested positive for SARS-CoV-2 infection, and notwithstanding some seemingly perinatal complications, the majority of neonates (296 out 298) born were negative for SARS-CoV-2 infection. Our observation is in agreement with those of other studies [**30-32]**, which implies that the consequences of SARS-CoV-2 infection on neurodevelopment were largely due to in utero effects rather than direct effects on the fetus. The reason for the low vertical transmission rate is that the placenta has low expression of the canonical receptors necessary for viral entry, which may explain the rarity of vertical transmission of SARS-CoV-2 [**32**]. Importantly, teratogenic effects of SARS-CoV-2 on the fetus were not observed, which is in contrast to other viral infections, such as Middle East respiratory syndrome coronavirus, CMV, herpes virus, Zika virus, and Rubella virus infections **[33-37]**.

In this study, we evaluated the infant’s neurobehavioral development 10-12 months post discharge, perhaps the adequate duration for effects to appear. In this study, overall, 69.1% of neonates at the end of 10-12 months showed normal developmental attainment, and only 10.1% (n=30) showed developmental delays. The PregCOV-19 living systematic review [**1**] found that approximately 95% of neonates born to SARS-CoV-2-positive mothers were reported to be born in good condition. However, it is not clear whether this systematic review included evaluations of neurodevelopmental delays in neonates. A study from the UK, in which information about educational and behavioral problems was collected from 177 children, found that 25% required support from a nonteaching assistant, 4% required a statement of special educational needs, and 3% were in a special school [**38**]. Many investigators have studied the effects of various viral infections, albeit not SARS-CoV-2, on pregnancy and fetal/neonatal outcomes and have reported that maternal influenza, hepatitis C, varicella-zoster, and other viruses produce distinct fetal brain structure and anatomy clinical pathology of varying severity **[39-42]**.

Of note, the role of cytokine storms, particularly the role of IL-6 in the pathogenesis of neurodevelopmental disorders, is not fully elucidated; however, in a longitudinal study, alterations in brain architecture, executive function, and working memory abilities were reported in 2-year-old neonates exposed to increased IL-6 levels during pregnancy [**43]**. An analysis of 214 patients with SARS-CoV-2 infection revealed that 36% had neurological symptoms **[44]**. Many case-control studies have demonstrated that elevated maternal cytokine levels affect the fetal brain, causing neurological disease **[45-47]**. The FIRS, due to increased IL-6 following maternal SARS-CoV-2 infection **[13]**, may induce a wide range of adverse neurodevelopmental sequelae, such as autism, psychosis, and neurosensory deficits, later in life [**48**], similar to certain bacterial infections [**49**]. However, long-term longitudinal studies are required to validate these associations.

Moreover, the risk of neurological effects on neonates differs by the trimester when SARS-CoV-2 infection occurred [**50**]. In this study, developmental delays were relatively more prevalent when infection occurred in the first and second trimesters than when infection occurred in the third trimester (P<0.001). Neonatal characteristics were similar between the groups. Moreover, infants born at less than 31 weeks gestation were more likely to have developmental delays than those born at more than 31 weeks gestation (10% versus 0.8%; P=0.002).

The findings of the study may be considered reliable, as the cohort of parents who participated in the study was well educated, and women were young (median age: 31 years). Three-fourths of women were devoid of pregnancy-induced hypertension or gestational diabetes.

## 5 Strength and Limitations

This is perhaps the first study reporting neurological assessments of neonates born to mothers with SARS-CoV-2-positive RT-PCR test results during pregnancy using the ASQ-3. The participating mothers were from the local area, which enabled good tracking for follow-ups. Participants were educated and responded to ASQ-3 questions appropriately. Thus, these points may be regarded as strength of the study.

This study also has a few limitations, the most important aspects of which are related to the sample size and methodology. For stronger conclusions, a larger and statistically valid sample size is required. Furthermore, the ASQ-3 were completed by the parents of the neonates; hence, biases in reporting cannot be ruled out. Nevertheless, this initial study may help healthcare authorities understand the need to assess the developmental attainment of infants born to mothers with SARS-CoV-2-positive test results during pregnancy. The ASQ-3, although a good screening tool, has limitations, and its results may not be strongly comparable to those of other tools, such as the Bayley III test; therefore, further larger studies with formal assessments of neurodevelopment are warranted.

## 6 Conclusions

This study was a hypothesis-generating study, and the findings based on a screening tool raise some concerns regarding neurodevelopmental effects on fetuses born to mothers with SARS-CoV-2-positive test results during pregnancy. A majority of the pregnant women had SARS-CoV-2 infections during their third trimester. Developmental delays were more prevalent when SARS-CoV-2 infection occurred in the first and second trimesters than when it occurred in the third trimester. Infants born at less than 31 weeks gestation were more likely to have developmental delays than those born at more than 31 weeks gestation. Further larger studies with formal developmental assessments are warranted to study the effect of SARS-CoV-2 on the developing brain.

## Data Availability

All data produced in the present study are available upon reasonable request to the authors

## Abbreviations

MIS-N: Multisystem inflammatory syndrome in neonates
SARS-CoV-2: Severe acute respiratory syndrome coronavirus-2
COVID-19: Coronavirus disease 2019
RT-PCR: Reverse transcriptase-polymerase chain reaction
CDC: Centers for Disease Control
CRP: C-reactive protein
IL: Interleukin
FIRS: Fetal inflammatory response

## Declarations

### Ethics approval and consent to participate

Ethics approval was granted by the Ministry of Health, (2021-1638), Government of Kuwait.

### Availability of data and materials

Not applicable

### Competing interests

The authors declare that they have no competing interests

### Funding

None

### Authors’ contributions

MA conceptualized and planned the study design, planned data collection, oversaw data collection, literature review, performed the text mining analysis, drafted and revised the final version of the manuscript. MA also performed the statistical analysis. AE, MK, MA, ZA, ZB, YB and HA conducted the literature review, contributed to the writing and reviewing of the manuscript. All other co-authors contributed to data collection and oversaw the manuscript.

## Acknowledgements

We would like to thank the respective register holders; Maternal and Perinatal COVID-19 Kuwait National Registry.

## Notes

### Competing Interest Statement

The authors have declared no competing interest.

### Funding Statement

This study did not receive any funding

### Author Declarations

Ethics approval was granted by the Ministry of Health, (2021-1638), Government of Kuwait.

